# Comparison of coil placement approaches targeting dorsolateral prefrontal cortex in depressed adolescents receiving repetitive transcranial magnetic stimulation: an electric field modeling study

**DOI:** 10.1101/2023.02.06.23285526

**Authors:** Zhi-De Deng, Pei L. Robins, Moritz Dannhauer, Laura M. Haugen, John D. Port, Paul E. Croarkin

## Abstract

**Background:** A promising treatment option for adolescents with treatment-resistant depression is high-frequency repetitive transcranial magnetic stimulation (rTMS) delivered to the left dorsolateral prefrontal cortex (L-DLPFC). Conventional coil placement strategies for rTMS in adults include the 5-cm rule, the Beam F3 method, and the magnetic resonance imaging (MRI) neuronavigation method. The purpose of this study was to compare the three targeting approaches to a computational E-field optimization coil placement method in depressed adolescents.

**Methods:** Ten consenting and assenting depressed adolescents (4 females, age: 15.9 ± 1.1) participated in an open-label rTMS treatment study. Participants were offered MRI-guided rTMS 5 times per week over 6–8 weeks. To compute the induced E-field, a head model was generated based on MRI images, and a figure-8 TMS coil (Neuronetics) was placed over the L-DLPFC using the four targeting approaches.

**Results:** Results show that there was a significant difference in the induced E-field at the L-DLPFC between the four targeting methods (*χ*^2^ = 24.7, *p <* 0.001). *Post hoc* pairwise comparisons show that there was a significant difference between any two of the targeting methods (Holm adjusted *p <* 0.05), with the 5-cm rule producing the weakest E-field (46.0 ± 17.4 V/m), followed by the F3 method (87.4 ± 35.4 V/m), followed by the MRI-guided (112.1 ± 14.6 V/m), and followed by the computationally optimized method (130.1 ± 18.1 V/m). The Bartlett test of homogeneity of variances show that there was a significant difference in sample variance between the groups (*K*^2^ = 8.0, *p <* 0.05), with F3 having the largest variance. In participants who completed the full course of treatment, the median E-field strength in the L-DLPFC was correlated with the change in depression severity (*r* = *–*0.77, *p <* 0.05).

**Conclusions:** The E-field models revealed inadequacies of scalp-based targeting methods compared to MRI-guidance. Computational optimization may further enhance E-field dose delivery to the treatment target.

## 1. Introduction

For adolescents with treatment-resistant depression (TRD), a safe, tolerable, and promising treatment is repetitive transcranial magnetic stimulation (rTMS) [1, 2, 3, 4]. The parameter space for dosing rTMS is vast and includes: stimulation target, coil targeting strategy, frequency, intensity, train duration, interstimulus intervals, pulses per session, number of sessions, and brain state. Of these parameters, a major area of interest is determining an optimal scalp coil placement for the left dorsolateral prefrontal cortex (L-DLPFC) [5]. Clinical studies on rTMS for adolescent depression have adopted coil placement approaches used in adults [1, 6, 7, 8]. However, adult coil placement approaches may not yield optimal dosing and clinical outcomes in adolescents. For instance, there are structural differences between adults and adolescents in head anatomy, including head size and myelination development, which can differentially affect the spread of induced electric field (E-field) in the brain. The rTMS induced E-field can be modeled in individual subjects using computational methods [9]. These computational models can enhance our understanding of the effect of neurodevelopmental variability and have utility in the individualization of rTMS dosing, such as coil placement.

The standard approach to placing a TMS coil over the L-DLPFC in adults is the “5-cm rule”, which involves measuring the scalp 5 cm along the parasagittal plane anterior to the activation hotspot in the motor cortex. While this method is easy to implement, it does not account for variations in head size, i.e., geodesic distances on scalp surface, between individuals. It has been shown that the 5-cm rule missed Brodmann area (BA) 9 in the L-DLPFC in more than two-thirds of adult subjects; the coil center often ended up in more dorsal regions such as BA6 or 8 [10]. Further, while adults’ head sizes remain relatively fixed, adolescents’ head circumference increases with age [11], resulting in a wider range of head sizes. Therefore, when applying the 5-cm rule in adolescents, there is an increased likelihood of missing the L-DLPFC target and hence producing more variability in the E-field dose and clinical outcomes. Another popular approach uses the International 10–20 electroencephalogram (EEG) system for positioning of TMS, in particular, the F3 site corresponded to parts of BA8, 9, and 46 within the L-DLPFC [12, 13]. Beam and colleagues developed an efficient way for locating the F3 position from a series of scalp measurements [14]. Compared to the 5-cm rule, the Beam F3 method better scales with head size, since the EEG electrodes placement system is based on measurements of head circumference, nasion–inion and tragus–tragus distances.

In addition to the scalp-based methods, another alternative is to use an MRI neuronavigated system to localize the L-DLPFC based on individual brain images. In our prior rTMS study for depressed adolescents, the 5-cm rule and the Beam F3 method yielded different locations that were on average 3.9 cm and 2.5 cm away from the MRI-derived L-DLPFC scalp target, respectively [7]. In adults, rTMS delivered with the MRI-based approach resulted in superior outcomes compared to the 5-cm rule, however, MRI acquisition is costly and therefore less practical for clinical TMS [15]. The Beam F3 method may be more cost-effective and practical for clinical TMS than using the 5-cm rule and the MRI-guided approaches [14, 15, 16]. Further, Mir-Moghtadaei and colleagues suggested that the Beam F3 method may provide a reasonable approximation than the MRI-guided neuronavigation on the L-DLPFC in a majority of adults [16].

In adolescents, little is known about the difference in induced E-field strength between three coil placements (5-cm rule, F3 method, MRI-based method). The goal of this study is to quantify and compare the induced E-field by three targeting approaches in a group of depressed adolescent participants who had previously undergone an acute course of 10 Hz rTMS delivered to the L-DLPFC. Further, we explore the utility of a computational approach [17] for optimizing individual coil placement to maximize the induced E-field in the L-DLPFC. A previous computational study using fast auxiliary dipole method showed that, compared to simply placing the coil above the center of mass of the target, the optimal scalp coil placement can be more than 10mm away, leading to an E-field strength of approximately 6% higher [18]. The Targeting and Analysis Pipeline (TAP) was subsequently developed that further demonstrated how a voxel-based ROI can be efficiently integrated with TMS coil placement optimization [17]. In this work, we build upon TAP and extend its capabilities to account for the size of the individual L-DLPFC masks. This work is an important initial step in part of a larger effort to improve rTMS protocols for the treatment of adolescent depression.

## 2. Method

### 2.1. Participants

This open-label rTMS study was conducted under an Investigational Device Exemption (#G110091) from the United States Food and Drug Administration and approved by the Mayo Clinic Institutional Review Board (ClinicalTrials.gov identifier: NCT01502033). The clinical outcomes of the trial have been previously published [6, 7]. Briefly, ten patients (4 females) with treatment-refractory major depressive disorder (MDD) between the ages of 13.9 and 17.4 years (mean ± standard deviation = 15.9 ± 1.1) participated in an open-label rTMS treatment study after providing informed consent and assent. Each patient met the Diagnostic and Statistical Manual of Mental Disorder, fourth edition, text revision (DSM-IV-TR), criteria for a major depressive episode based on a semi-structured diagnostic interview, the Schedule for Affective Disorders and Schizophrenia for School-Age Children-Present and Lifetime version (K-SADS-PL) [19]. Participants had moderate-to-severe symptom severity ratings as evidenced by a baseline Children’s Depression Rating Scale-Revised (CDRS-R) [20]. A total CDRS-R score of 40 or greater at baseline was required for inclusion criteria. Further, all patients had to have at least one prior failed antidepressant medication trial as defined by the Antidepressant Treatment History Form [21].

### 2.2. Motor hotspot and resting motor threshold determination

Motor hotspot and motor threshold were determined from visual observation of movement in the abductor pollicis brevis (APB) muscle of the right hand during TMS over the contralateral motor cortex using the NeuroStar coil (Neuronetics, Inc., Malvern, PA). MT was measured in units of Standard Motor Threshold (SMT). One SMT is the output setting that corresponds to an induced E-field of 135 V*/*m at a point located 2 cm along the central axis of the treatment coil from the surface of the scalp into the patient’s cortex. This corresponds to the average motor threshold level observed in a large adult population [22]. We converted the SMT unit to the rate of change of coil current, d*I/*d*t*, which is the input to the E-field simulations (see conversion in Supplementary Material).

### 2.3. Imaging

Patients were custom fitted with a swim cap on which the APB motor hotspot, 5-cm site, F3 site, mid-frontal, left mastoid, right mastoid, right parietal, and right frontal were marked with fiducial markers. T1-weighted structural MRI data was acquired on a GE 3-T DV750 scanner equipped with an eight-channel head coil (true-axial fast 3D-SPGR sequence, repetition time (TR) = 12.6 ms, echo time (TE) = 5.6 ms, flip angle = 15 degrees, voxel dimensions = 0.49 × 0.49 × 1.5 mm^3^, field of view = 250 × 250 mm^2^, slice = 1.5 mm, matrix = 512 × 250 pixels).

### 2.4. Treatment intervention

The treatment target was derived from the T1-weighted anatomical images using the Medtronic StealthStation™ navigation system (Medtronic Navigation, Inc., Louisville, CO) by creating a real-time surgical navigation on patients’ radiological images. The DLPFC brain target (DBT) was defined as a 20 × 20 × 20 mm^3^ voxel in the L-DLPFC according to the following anatomical guidelines [7]: 1) the “inferior plane” of the corpus callosum was identified as a line that abutted the inferior margins of the rostrum and splenium of the corpus callosum; 2) a 20-mm thick coronal-oblique localizer slice was acquired perpendicular to the inferior plane, such that the center of the localizer slice was placed 10-mm anterior to the genu of the corpus callosum, and the posterior edge of the slice abutted the anterior margin of the rostrum of the corpus callosum; 3) the deepest portion of the superior frontal sulcus was identified to be the DBT. The averaged center of the DBT voxels was projected through the shortest straight path to the scalp, yielding the coordinate for the DLPFC scalp target (DST). Over the DST, 10 Hz rTMS was delivered using the NeuroStar Therapy System, at an intensity of 120% SMT, 5 days per week, over 6–8 weeks, up to a total of 30 sessions.

### 2.5. DLPFC gray matter voxel mask

The L-DLPFC mask was binarized (MRIcroGL, https://www.nitrc.org/projects/mricrogl/) for the skull-stripped MNI-152 template using the following procedures [23, 24]: 1) measure the distance between the most anterior point of the frontal pole and the most anterior ipsilateral temporal pole (*d*_FP–TP_); 2) measure the distance from the tip of the temporal pole anteriorly 20% *d*_FP–TP_ to mark the posterior vertical boundary of L-DLPFC; 3) measure the distance from the tip of the frontal pole posteriorly 40% *d*_FP–TP_ to mark the anterior vertical boundary of L-DLPFC; 4) measure the distance from the most inferior part of the temporal lobe to the most superior part of the brain (*d*_BB–TB_); 5) in the coronal slice measure the distance from 50% *d*_BB–TB_ superiorly to mark the inferior boundary of the superior frontal sulcus The L-DLPFC mask in MNI space was transformed into the subject’s space using the FLIRT (FMRIB’s Linear Image Registration Tool) function in FSL with 12 degrees of freedom [25].

### 2.6. E-field modeling

The fiducial markers on the T1-weighted anatomical images were manually erased in ImageJ [26]. To simulate the E-fields, T1-weighted images were segmented (`mri2mesh` pipeline) into six tissue compartments using SimNIBS 3.0 [27]: skin, skull, cerebrospinal fluid, gray matter, white matter, and eyes, with assigned isotropic conductivities of 0.465 S*/*m, 0.010 S*/*m, 1.654 S*/*m, 0.275 S*/*m, 0.126 S*/*m, and 0.5 S*/*m, respectively. For one of the patients, the `mri2mesh` pipeline errored out in SimNIBS 3.0 but could successfully processed with SimNIBS 2. The segmentation was meshed as a tetrahedral finite element model of the participants’ head, where the Neuronetics coil was centered at the 5-cm, F3, and MRI-derived sites, oriented 45 degrees toward midline. The E-field was simulated for the rate of change of the coil current, d*I/*d*t*, corresponding to individual treatment dose in SMT units.

For coil placement optimization, we used a direct solver implemented in SimNIBS since there was no dipole coil file (ccd file) available for the Neuronetics coil. Using the TAP software [17], we registered the L-DLPFC gray matter voxel mask to the subject and determine the ROI center voxel coordinate and size as represented by a sphere. The average spherical radius of the individual ROIs is 11.4 ± 0.48 mm. The E-field optimization was a discrete search, for which different coil centers (on a 1-mm grid) and orientations (4 degree increments) are evaluated regarding the maximal averaged E-field magnitude within a 20-mm radius around the scalp-projected point of the L-DLPFC center.

The SimNIBS `msh2nii` command line tool was used to interpolate simulated E-field magnitude values for gray matter MRI voxels within the L-DLPFC mask. The median E-field strength was extracted from the L-DLPFC gray matter voxel mask. The median E-field values from the four targeting methods were compared using the Friedman test followed by *post hoc* pairwise Wilcoxon tests with Holm adjustment. Finally, a recent report suggested that the normal component of the TMS-induced E-field is correlated with depressive symptom relief in treatment-resistant depression in adults [28]. Thus, we explored the relationships between median magnitude of the E-field and its normal component with the change in CDRS for participants who have completed a full course of treatment. The E-field normal component was calculated by mapping the vectorized E-field and mask mesh files to the cortical surface (Freesurfer’s FsAverage surface) with the `msh2cortex` command line tool, which utilized the superconvergent path recovery method [29].

## 3. Results

Of the ten participants enrolled in the study, one (Participant #3 in Figure 1) had an unusually high motor threshold (1.54 SMT). This high threshold resulted in high treatment stimulation intensity (1.85 SMT) and high median L-DLPFC E-field (214.0 V*/*m). The high stimulation intensity contributed to scalp discomfort that the participant was unable to tolerate; the participant dropped out of the study after the first rTMS session. The subsequent E-field analysis was performed with the data from the remaining nine participants.

**Figure 1:**
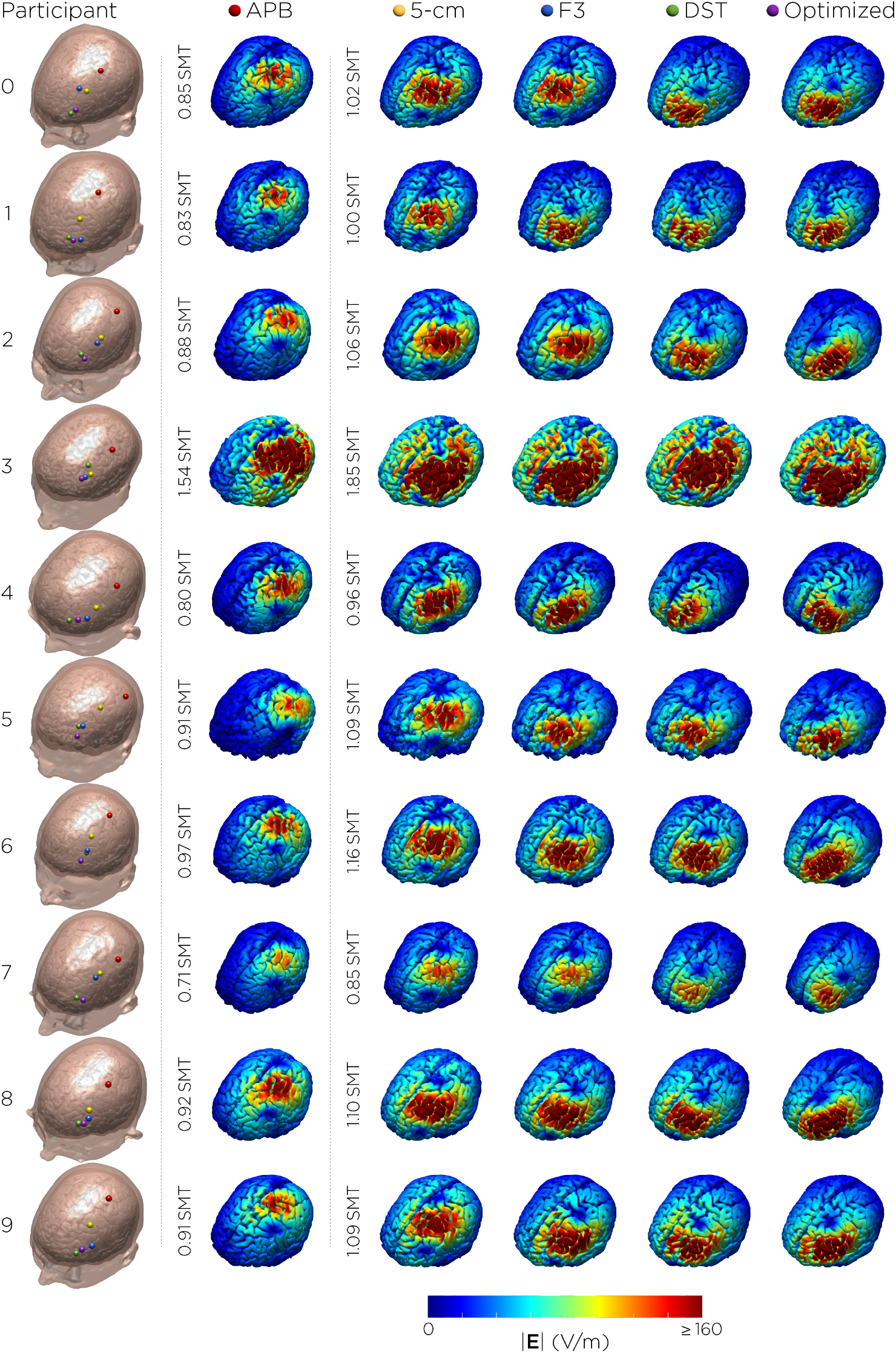
TMS targets and corresponding E-field distribution on the brain. The E-fields for the APB motor hotspot (red dot) stimulation were simulated with input intensities corresponding to individual motor thresholds (SMT units). The E-fields for the treatment targets [5-cm rule (yellow), Beam F3 (blue), MRI-derived DLPFC Scalp Target (green), and computationally optimized coil placement (purple)] were simulated at 120% motor threshold. Participant #3 dropped out from the study after the first session due to unusually high motor threshold and treatment stimulation intensity.

There was a significant difference in the induced E-field at the L-DLPFC between the four targeting methods (*χ*^2^ = 24.7, *p <* 0.001). *Post hoc* pairwise comparisons showed that there was a significant difference between any two of the targeting methods (Holm adjusted *p <* 0.05), with the 5-cm rule producing the weakest E-field (46.0 ± 17.4 V*/*m), followed by the F3 method (87.4 ± 35.4 V*/*m), followed by the MRI-guided DST method (112.1 ± 14.6 V*/*m), and followed by the computationally optimized method (130.1 ± 18.1 V*/*m). The Bartlett test of homogeneity of variances showed that there was a significant difference in sample variance between the groups (*K*^2^ = 8.0, *p <* 0.05), with the F3 method having the largest variance in E-field strengths (Figure 2).

**Figure 2:**
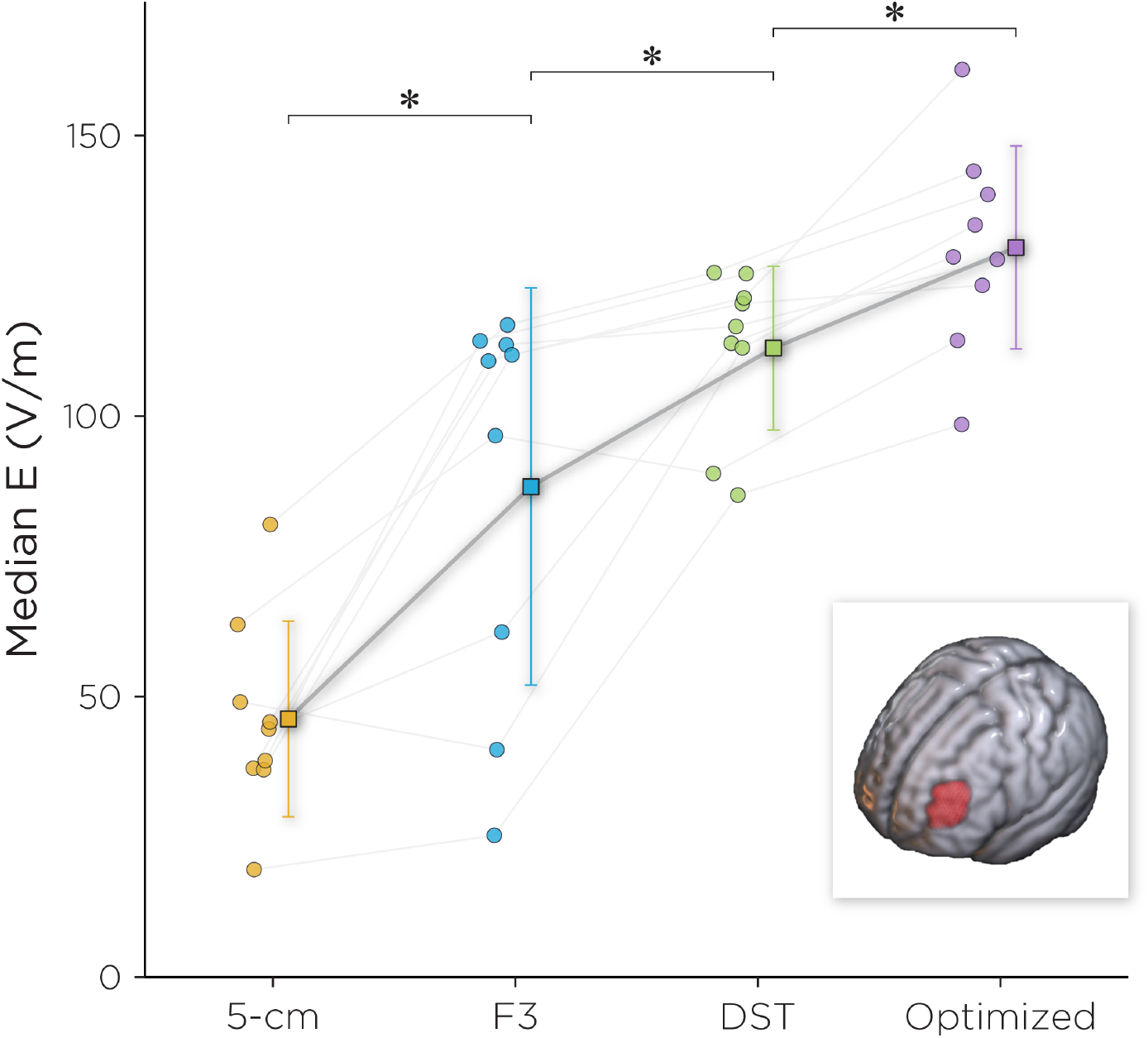
Median E-field for the four targeting strategies. *Post hoc* pairwise comparisons showed that there was a significant difference between any two of the targeting methods (Holm adjusted *p <* 0.05). The inset shows L-DLPFC voxel mask on which the median E-field was extracted.

Figure 3 explores the relationship between the L-DLPFC magnitudes of the E-field and its normal component with the change in CDRS scores. In addition to participant #3 noted above, two other participants did not complete the full course of rTMS treatments due to worsening depression and anxiety; their data were removed from the correlational analysis with E-field and clinical outcomes. The median E-field magnitude was linearly correlated with the change in CDRS (*r* = *–*0.77, *p <* 0.05), while the normal component of the E-field did not show such relationship.

**Figure 3:**
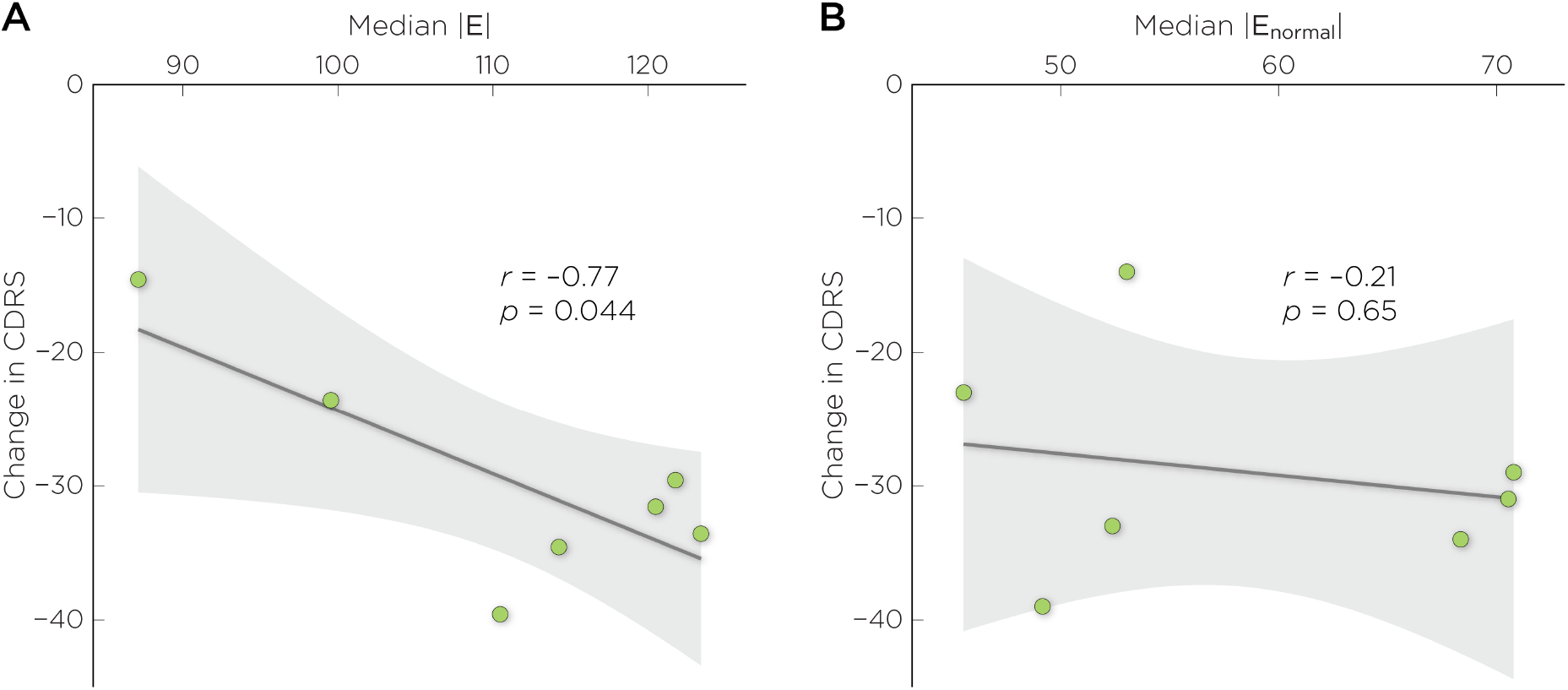
Correlation between the change in CDRS with A) magnitude of the E-field, and B) magnitude of the normal component of the E-field.

## 4. Discussion

Analogous to Global Positioning System (GPS) navigation, there are two key components to TMS targeting: first is to determine the location of the destination address (target identification); second is to chart the path to the destination accurately and precisely (coil placement). The search for the optimal target for rTMS depression treatment has generated much recent research interest. The DLPFC is the standard targeting site used in rTMS studies for both adults and adolescents. The conventional rationale to target the DLPFC is based on its hemispheric asymmetry and imbalance of activities in depression, in which the left DLPFC is hypoactive and the right DLPFC is hyperactive [30, 31]. To balance both hemispheres of the DLPFC, high-frequency rTMS (10–20 Hz) is used to increase neuronal excitability of the L-DLPFC while low-frequency rTMS (1 Hz) is used to induce neuronal inhibitory of the R-DLPFC [32]. Other approaches to identify a target within the DLPFC use functional neuroimaging, for example, using resting-state fMRI to determine the location of the maximum anticorrelation between the L-DLPFC and the subgenual anterior cingulate cortex (sACC) [33, 34], or task-based fMRI to determine the location of the peak activation in engagement with a goal priming task [35]. Another approach attempts to engage the DLPFC node in the frontal–vagal pathway that overlaps with functional nodes of the depression network [36]. The resultant Neuro-Cardiac-Guided TMS (NCG-TMS) technique uses TMS-induced heart rate deceleration as a marker for target engagement and stimulation site determination [37]. Finally, although the DLPFC has been the most popular targeting site, recent studies have attempted to target other regions that are involved in emotion regulation [38], such as the dorsomedial prefrontal cortex [39, 40, 41] and the right orbitofrontal [42].

Once the treatment target has been identified, the other aspect of TMS targeting is to choose a navigation strategy that would best locate the stimulation site. The clinical standard for targeting the L-DLPFC has been the 5-cm rule. In this work, we showed that among targeting strategies, the 5-cm rule produces the lowest E-field in the L-DLPFC. In view of the dose–response function between E-field strength and antidepressant outcome (Figure 3A), the consistent underdosing of the E-field by the 5-cm rule could lead to suboptimal therapeutic effect. For example, in a blinded, randomized, sham-controlled clinical study on the effectiveness of high-frequency rTMS for TRD in adolescents, the 5-cm rule was used to target the L-DLPFC. The trial found no statistically significant difference between the active and sham groups in antidepressant efficacy [43]. There have been proposed variations to the 5-cm rule to improve its accuracy, such as the 5.5-cm rule [44] and the “5 cm + 1 cm” (6-cm) rule [45]. Johnson and his colleagues compared the targeting variations using the 5-cm rule and the 6-cm rule [45]. They found that both targeting strategies produced similar intra-variability across adult subjects, in which the 6-cm rule would shift the variability range more anterior. By using the 6-cm rule, they proposed that this variation could offer more therapeutic effects. However, the authors did not compare the clinical efficacy between these targeting variations [45]. An additional limitation of the 5-cm rule concerns the motor hotspot and threshold determination technique, which is often done with observed movement of the target muscle method (OM-MT) without electromyography (EMG) recording. Prior work suggests that the OM-MT method yields significantly higher MTs compared to thresholding method based on EMG [46]. The OM-MT method can produce suboptimal motor hotspot to which the 5-cm target is anchored to, and elevated risk due to overestimation of the MT to which the treatment stimulation intensity is scaled to.

The Beam F3 method is another navigation strategy to target the L-DLPFC. In our E-field modeling, we showed that compared to the 5-cm rule, the Beam F3 method can achieve more accurate targeting, but not necessarily more precise. That is, on average, the Beam F3 method produces higher E-field in L-DLPFC, but in our sample, showed more variability across individuals. A recently published report showed that in adults, when comparing coil distance from BA46, the Beam F3 method produced the largest variance from the target compared to the 5-cm rule, and the MRI-guided approach [47]. There have been proposed modifications to the Beam F3 method. For example, one study modified the method to better estimate the optimized anti-subgenual TMS target [48], using the MNI anatomical template to determine the distances (measurements that would be required) from anatomical landmarks to the target average coordinate that showed the greatest DLPFC–sACC anticorrelation. This variation showed that the newer anterior L-DLPFC estimate was 21.5 ± 1.4 mm more inferior-posterior to F3, while the posterior L-DLPFC is 37.0 ± 0.6 mm more posterior to F3.

Neuronavigation is another targeting strategy with different adaptations. Structural MRI-guided methods of localizing the DLPFC can be based on targeting specific Brodmann area sites, or using other anatomical definitions. The DLPFC comprises two different cyto-architectural sub-regions: BA9 and 46. A preliminary study randomly assigned participants to two treatment groups to receive either rTMS over BA9 or over BA46, stimulation of both Brodmann areas led to similar antidepressant responses [49]. Other studies target a site in the junction of BA9 and 46, using coordinates in a standard atlas space, such as the Talairach atlas [50]. Our study defined the L-DLPFC as a point projection of a voxel in the brain based on a series of anatomical guidelines [6, 7], as described in the Methods. Functional MRI-guided targets can be derived from group-averaged functional maps [51, 52, 53, 54] or from individualized connectivity [55, 56]. The optimal treatment target can vary across individuals; another factor that contributes to variability is interindividual differences in head anatomy, which can influence the spatial distribution of the induced E-field [57]. In this work, we showed that given a target of interest, computational optimization can be used maximize the E-field delivery to the target. More sophisticated algorithms have been proposed to combine individual functional connectivity patterns, and E-field optimization, to determine coil placements that would not only maximize local stimulation, but also account for downstream effects of TMS, i.e., to maximize brain network engagement [58, 59, 60, 61]. Systematic clinical trials are needed to prospectively compare the antidepressant efficacy of these strategies in adults and adolescents.

There are a number of limitations to consider for the interpretation of this work. First, this is a pilot study with small sample size. This is compound by drop out–several participants were unable to complete the full treatment course. Although data from the remaining participants did suggest a dose–response relationship (Figure 3A), it is to be interpreted with caution. The second limitation concerns the head model tissue properties used in the E-field simulations, particularly in developing adolescents. During adolescence, the brain undergoes significant changes in terms of both structure and function. One of the key changes that occurs during this time is the continuation of myelination that helps to refine and optimize the connectivity between different brain regions. Studies have shown that myelination in adolescents is lower—particularly in the frontal lobes—compared to adults [62]. Differences in the degree of myelination may in part explain age differences in motor threshold, which is higher in younger individuals [63]. When modeling E-fields in the head and brain, it is common to assign standard isotropic conductivity values for different tissues based on values that have been reported in the literature; this tissue properties assignment does not factor in the effects of age. Finally, our study does not address the effects of broad stimulation of the DLPFC with nonfocal TMS coils.

## 5. Conclusion

Proper placement of the TMS coil is necessary to ensure that the desired brain region is adequately stimulated, while minimizing the risk of stimulating unintended areas. Understanding the factors that influence delivered E-field dose in the brain can help to improve the effectiveness of rTMS as a treatment. In this computational study, we modeled a group of adolescents receiving rTMS targeted to the L-DLPFC. We made within-subject comparisons of three targeting strategies: the 5-cm rule, the Beam F3 method, and MRI-guided targeting. These models showed various shortcomings of the scalp-based targeting methods: the 5-cm rule underdosed E-field to the L-DLPFC, and the Beam F3 method exhibited higher inter-subject variability. In the study participants who received a full course of MRI-guided rTMS, the data suggested a dose–response relationship between the induced E-field and clinical improvement. This motivates the use of computational techniques to further optimize E-field delivery to the treatment target.

## Data Availability

All data produced in the present study are available upon reasonable request to the authors

## Acknowledgments

Z.-D. Deng, P. Robins, and M. Dannhauer are supported by the National Institute of Mental Health (NIMH) Intramural Research Program (ZIAMH002955). P. Croarkin is supported by the NIMH R01MH113700 grant: “Glutamatergic and GABAergic Biomarkers in TMS for Adolescent Depression.” The study was also supported by the O’Shaughnessy (Woolls’) Foundation. Neuronetics provided equipment support and Senstar shields through an investigator-initiated trial program to Dr. Croarkin. This work utilized the computational resources of the NIH HPC Biowulf cluster (http://hpc.nih.gov).

## Supplementary material

### S1. Estimation of d*I/*d*t* for Standard Motor Threshold

The output level units on the NeuroStar System are Standard Motor Threshold (SMT units). 1 SMT is the output setting that corresponds to an induced electric field of 135 V*/*m at a point located 2.0 cm along the central axis of the treatment coil from the surface of the scalp into the patient’s cortex. This corresponds to the average motor threshold level observed in a large patient population [1].

**Figure S1.**
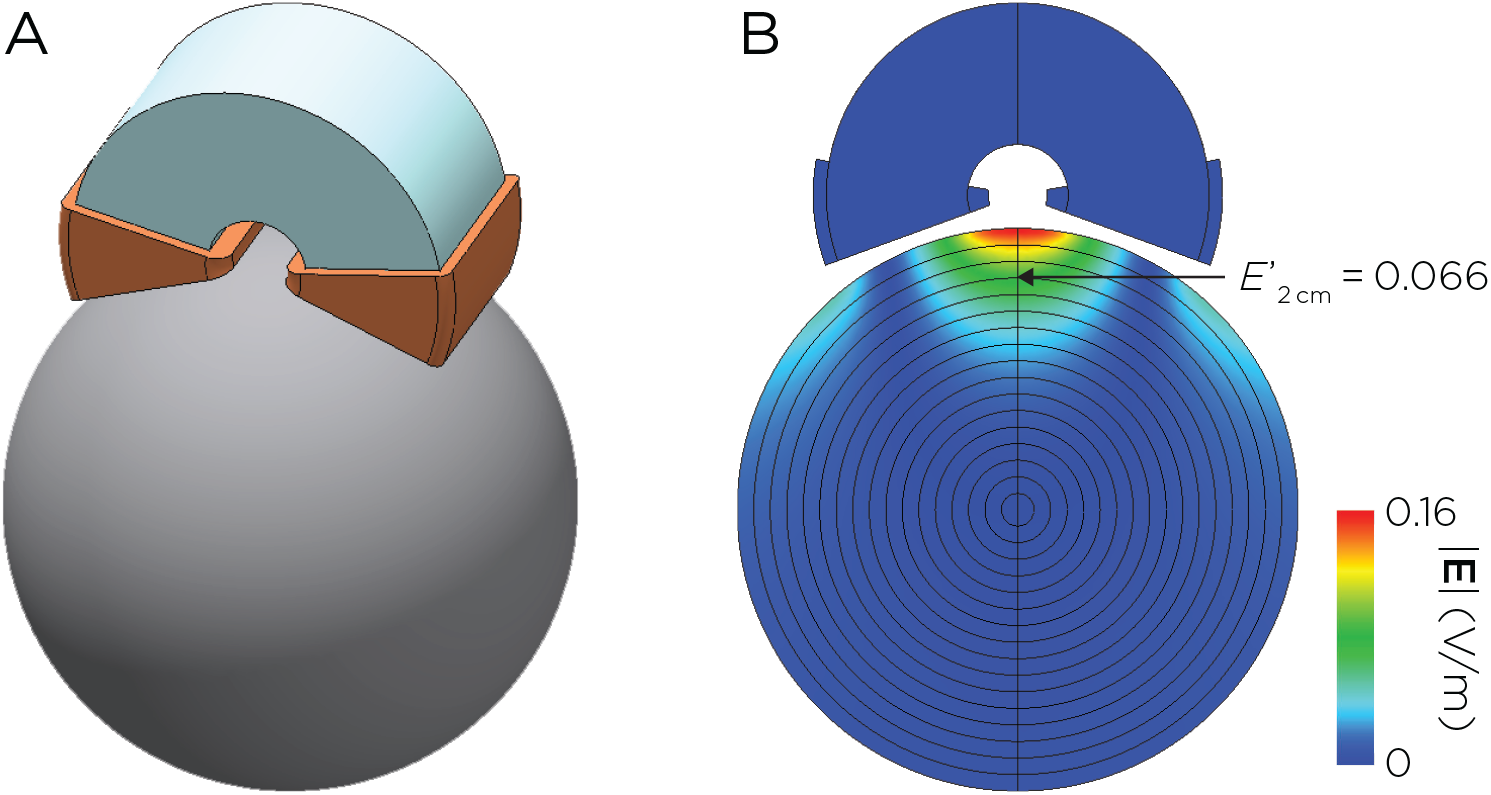
A. Model of the Neuronetics coil with C-shaped ferromagnetic core. B. Induced E-field distribution in a spherical head model with unit coil current *I*_0_ = 1 A and frequency *ω*_0_ = 2*π* × 5 kHz.

In order to estimate the d*I/*d*t* input for the individual E-field simulations, we first computed the E-field in a spherical head model. The head was modeled as a homogeneous sphere with a radius of 8.5 cm and isotropic conductivity of 0.33 S*/*m. The Neuronetics coil consisted of a figure-8 winding with a C-shaped ferromagnetic core [2, 3] (Figure S1A). The spatial E-field distribution was computed for an arbitrary coil current *I*_0_ = 1 A and frequency *ω*_0_ = 2*π* × 5 kHz using the MagNet Time Harmonic solver, yielding a E-field value at 2-cm depth of 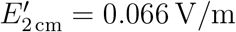 (Figure S1B). d*I/*d*t* was subsequently calculated for the desired field strength of 135 V*/*m at 2-cm depth:

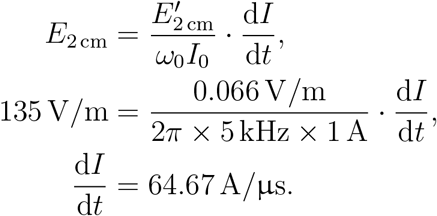

Using this d*I/*d*t* value, the mean maximum induced E-field at the motor cortex of the adolescent subjects at individual motor threshold is 141.3 ± 14.7 V*/*m, which is comparable to the reference SMT field strength of 135 V*/*m reported for adults (*t* = 1.30, *p* = 0.23).

### S2. Coordinate transformation

For the pilot clinical trial, coil targets were marked using the Medtronic StealthStation™ S7 equipped with an AxiEM frameless localization system running Synergy Cranial 2.2.6. The surgical navigation software has a proprietary coordinate system that needed to be transformed into MRI coordinates.

**Figure S2.**
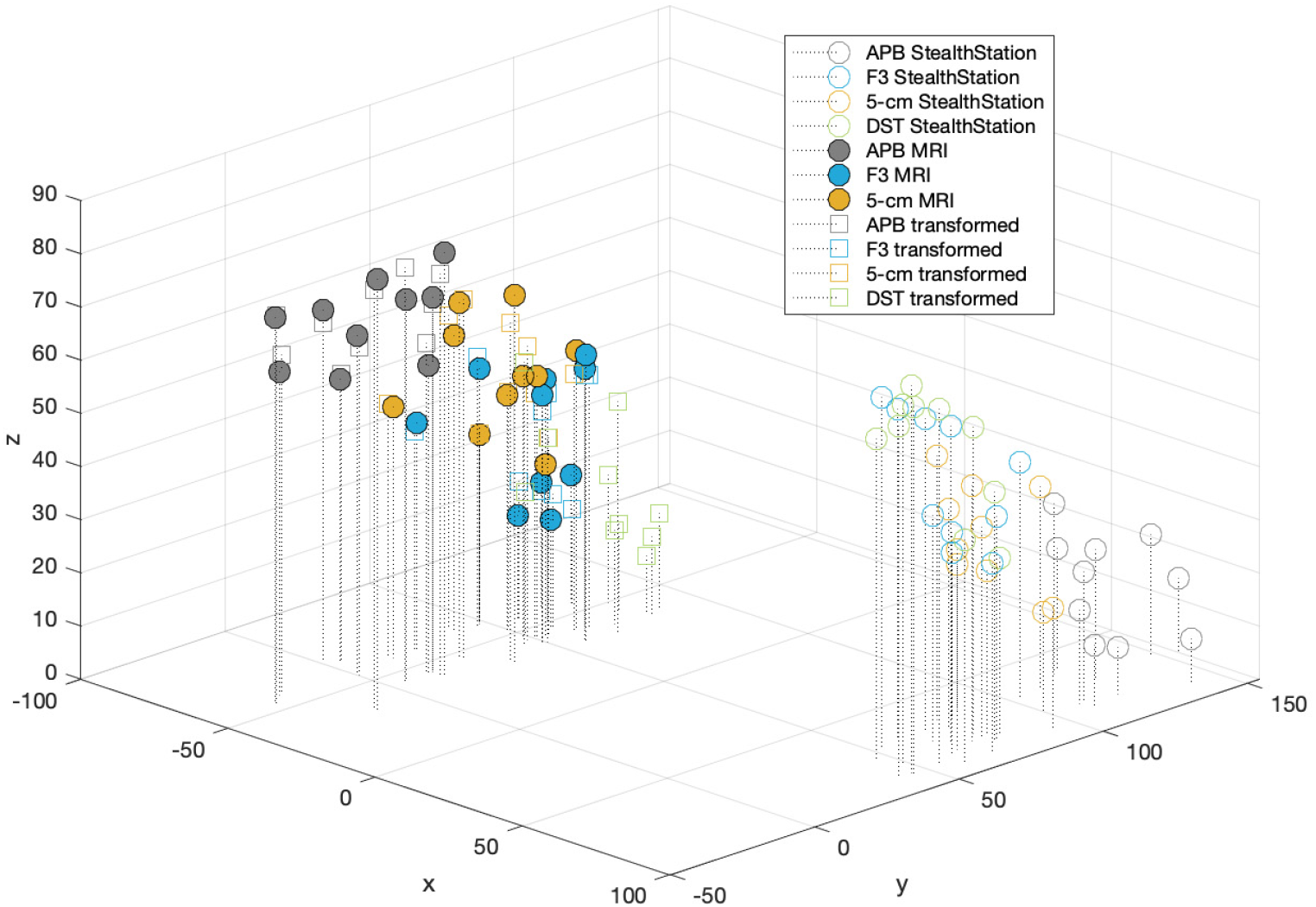
Individual scalp targets in StealthStation coordinates (empty circles), MRI fiducial markers (filled circles), and the transformed target coordinates in MRI space (squares).

We first performed 3D rendering the individual MRIs in MATLAB and manually marked the center of the fiducials (APB, 5-cm, and F3 targets) on the scalp. The ensemble of target coordinates from all ten subjects were used to derive the affine transformation from the surgical navigation system to MRI space. The DLPFC Scalp Target (DST) for individual subjects were subsequently transformed to MRI space for the E-field simulations. Figure S2 shows the individual scalp targets in StealthStation coordinates (circles), MRI fiducial markers (stars), and the StealthStation-to-MRI transformed coordinates (squares). The error between the manual MRI fiducial markers and the final transformed target coordinates is less than 2 mm in the *x* and *y* directions, and less than 6 mm in the *z* direction.

## Notes

### Competing Interest Statement

The authors have declared no competing interest.

### Clinical Trial

NCT01502033

### Author Declarations

This open-label rTMS study was conducted under an Investigational Device Exemption (#G110091) from the United States Food and Drug Administration and approved by the Mayo Clinic Institutional Review Board.

